# Mental health-related hospitalisations among adolescents with previous child protection contact from birth to age 11

**DOI:** 10.1101/2023.09.19.23295224

**Authors:** Jessica Judd, Rhiannon Pilkington, Catia Malvaso, Alexandra Procter, Alicia Montgomerie, Jemma Anderson, Jon Jureidini, Julie Petersen, John Lynch, Catherine Chittleborough

**Affiliations:** School of Public Health, BetterStart Health and Development Research, University of Adelaide, Adelaide, Australia; School of Psychology, University of Adelaide, Adelaide, Australia; Robinson Research Institute, University of Adelaide, Adelaide, Australia; Department of General Adolescent Medicine, Women’s and Children’s Hospital; Adelaide Medical School, University of Adelaide, Adelaide, South Australia; Department for Child Protection, Adelaide, South Australia; Population Health Sciences, University of Bristol, Bristol, United Kingdom

**Author notes:** Corresponding author: Jessica Judd, Address: School of Public Health, University of Adelaide, Level 4, Rundle Mall Plaza, 50 Rundle Mall, South Australia, 5005. Co-senior authors (John Lynch, Catherine Chittleborough).

## Abstract

**Objectives:** To examine the burden of mental health-related hospitalisations among adolescents by levels of previous child protection contact.

**Design, setting and participants:** Whole-of-population study of children born in South Australia, 1991-1999 (n=175,115), using de-identified linked administrative data from the Better Evidence Better Outcomes Linked Data (BEBOLD) platform.

**Main outcome measures:** Adolescents: proportion of adolescents aged 12-17 years with mental health hospitalisations; Hospitalisations: proportion of all adolescent mental health hospitalisations according to the level of child protection contact from 0-11 years.

**Results:** Overall, 15.5% (27,203/175,115 children) of adolescents had a history of child protection contact between ages 0-11 years, and 3.2% (5,646/175,115; 95% CI, 3.1 – 3.3) had a mental health-related hospitalisation between ages 12-17 years. Of the 10,633 mental health-related hospitalisations, 44.9% (95% CI, 44.0 – 45.9) were among adolescents with previous child protection contact even though they comprised only 15.5% of the study population. Of 5,646 adolescents with at least one mental health-related hospitalisation, 40.4% (95% CI, 39.1 – 41.7) had previous child protection contact. Among the population who experienced out-of-home care, 17.5% (209/1,191; 95% CI, 15.5 – 19.8) had experienced a mental health-related hospitalisation during adolescence, compared to 2.3% (3,366/147,912; 95% CI, 2.2 – 2.4) of adolescents with no prior child protection contact.

**Conclusion:** Almost 45% of mental health hospitalisations for 12-17-year-olds occurred among children who had child protection contact, despite that group comprising only 15.5% of the study population. Potential trauma sequelae associated with child protection history is important to consider in the response to adolescents hospitalised due to mental health challenges.

**Significance of Study:** *The known:* Adolescent mental health is an important public health issue and those in child protection are at higher risk of experiencing mental health challenges.

*The new:* We have quantified the burden of adolescent mental health hospitalisations attributable to the population with prior child protection system contact. For adolescents aged 12-17 years, those with a child protection history accounted for 44.9% of all adolescent mental health hospitalisations.

*The implications:* Potential trauma sequelae associated with child protection history are important to consider in the response to adolescents hospitalised due to mental health challenges.

## Introduction

Mental health challenges among adolescents are a major public health issue. For instance, suicide is the leading cause of death among 15-24-year-olds in Australia.^1^ Parent-or carer-reported estimates of “mental health disorders” from the 2013-14 Australian Child and Adolescent Survey of Mental Health and Wellbeing indicated that 14.4% of 12-17-year-olds had a “mental health disorder” in the previous 12 months.^2^ In the 2021 South Australian Population Health Survey, 19.7% of those aged 10-15, and 35.3% of adolescents aged 16-17 reported being told by a doctor that they had a “mental health condition” in the last 12-months.^3^ The COVID-19 pandemic may have exacerbated these mental health challenges.^4, 5^

The scale of mental health challenges means that services for young people may be overwhelmed and under-resourced in Australia.^6^ The current primary care model only meets the mental health needs of a minority of young people, with many requiring more comprehensive and multidisciplinary approaches due to complex needs.^7^ This has a flow-on effect in the health system as adolescents may present to hospital for mental health challenges when other services are unavailable.^6^ The rise in mental health-related hospitalisations for adolescents could therefore be considered to relate to a lack of available care and social supports or simply as one relevant indicator of adolescent mental health. Between 2010-11 and 2020-21, overnight mental health-related hospitalisation rates nearly doubled from 35 to 62 hospitalisations per 10,000 population for those aged 12-17.^8^

Teicher et al. considers child maltreatment to be “the most important preventable risk factor for psychiatric disorders” based on population attributable fractions for a range of psychopathology types ranging from 30% to 67%.^9^ We know child protection system contact and child maltreatment is common. In Australia in 2020-21, there were 531,900 notifications of alleged maltreatment^10^; data from South Australia has shown 1 in 4 children have been the subject of a child protection notification by age 10; ^11^ and in New South Wales 13.8% of children were notified to child protection services by age 5.^12^ Previous research has provided some evidence of the association between child protection system contact, child maltreatment and mental health hospitalisations. In a South Australian study that described frequency and reasons for hospitalisation by lifetime child protection involvement, mental health was amongst the top three reasons for hospitalisation for 13-17-year-olds who had experienced out-of-home care but not for those who had no child protection system contact.^13^ A New South Wales study found children who were the subject of a report to child protection services by age 6 were more than twice as likely to be diagnosed with a mental health disorder during middle childhood (6-14 years) compared to children who had never had child protection system contact.^14^ Another study in Western Australia found that 20% of children with substantiated maltreatment had a mental health diagnosis compared to 3.6% of children with no child protection contact.^15^

There is an opportunity to understand how the association between child protection system contact and mental health hospitalisations impacts the hospital system. Our study has taken the perspective of the hospital system and focused on investigating the burden of mental health-related hospitalisations that come from adolescents with a history of child protection contact. This perspective is relevant to clinicians trying to understand the background characteristics of adolescents presenting to hospital for mental health reasons.

Our study used linked whole-of-population data to investigate the burden of mental health-related hospitalisations among adolescents aged 12-17 years in South Australia who experienced child protection system contact from ages 0-11. We first described the overlap in the populations of children with child protection contact up to age 11 and those who had experienced a mental health-related hospitalisation during adolescence from age 12-17. We then quantified the number of individuals with a mental health-related hospitalisation (person view) and the number of mental health-related hospitalisations (a hospital view), to examine the burden of mental health-related hospitalisations by levels of prior child protection system contact among adolescents with at least one hospitalisation.

## Methods

### Data Sources

The Better Evidence Better Outcomes Linked Data (BEBOLD) platform contains de-identified whole-population linked administrative data on all South Australian children born from 1991 onwards. Data were probabilistically linked by an independent agency using demographic characteristics. Australian data linkage systems typically estimate a false linkage rate of 0.1^16^-0.5%.^17^ We used data from the South Australian Department for Child Protection, Admitted Patient Care (hospital data) and the South Australian Perinatal Statistics collection from the Department for Health and Wellbeing.

Ethics approval was granted by the South Australian Department of Health Human Research Ethics Committee (2022/HRE00137), the University of Adelaide HREC (H-185-2011), and the Aboriginal Health Research Ethics Committee (REC2411/9/14). Approval to use these data was also provided by the custodians of each data source.

### Study Population

Nine birth cohorts born in South Australia between 1^st^ January 1991 and 31^st^ December 1999 (n=175,115) were followed from ages 0-11 years for child protection contact, and then from ages 12-17 for mental health-related hospitalisations.

### Mental Health Hospitalisations

Mental health-related hospitalisations were defined using International Statistical Classification of Diseases and Related Health Problems, 10th Revision, Australian Modification (ICD-10-AM) codes (Supplementary Table 1) in primary or additional diagnoses. All external cause codes for self-harm were also included. These codes include those used by the Australian Institute of Health and Welfare and additional codes recognised as mental health hospitalisations in other Australian studies.^14, 15, 18, 19^

### Child Protection System Contact

Children were classified into the highest level of contact they had with the child protection system at ages 0-11 years (Box 1): no contact, notification but not screened in, screened-in notification but not investigated, investigation but not substantiated, substantiation and out-of-home care (including care and protection orders).

##### Box 1: Definitions of Child Protection Contact Levels^25^

| Level of Child Protection Contact | Definition |
| --- | --- |
| Notification | A report to the Child Protection agency concerning suspected child abuse or neglect. |
| Screened in | A term used to indicate whether the notification is of sufficient concern to warrant intervention by the Child Protection Agency. Those that meet the threshold are screened in; those that do not are screened out. |
| Investigation | A determination of whether an incident of child abuse or neglect has occurred, and the circumstances of its occurrence. |
| Substantiation | A professional judgement as to whether abuse or neglect has occurred. |
| Out-of-home-care (OOHC) | System of caring for a child who is removed from their family of origin; includes (but is not limited to) family-based care, emergency care and residential care. |

### Data Analysis

We calculated the proportion of adolescents who had at least one mental health-related hospitalisation, and for specific mental health diagnoses (between ages 12-17) by highest level of child protection contact (ages 0-11). We looked at where the patient was referred or transferred from for each mental health-related hospitalisation as well as number of recurrent hospitalisations for adolescents with at least one mental health-related hospitalisation between 12-17 years. For the analysis of the mental health burden from the perspective of the hospital system, we included only those adolescents with at least one hospitalisation at ages 12-17 years. First, we examined the burden of mental health-related hospitalisations among adolescents with prior child protection system contact. For this, we examined the distribution of mental health-related hospitalisations at ages 12-17 years, both in terms of the number of individuals with at least one mental health-related hospitalisation (person view) and the number of mental health-related hospitalisations (hospital view), by the highest level of child protection system contact experienced between ages 0-11 years. Second, we examined the proportion of all individuals hospitalised (person view), and the proportion of hospitalisations (hospital view), for 12-17-year-olds for all causes that were due to mental health by highest level of child protection contact and age.

Analyses were undertaken using STATA/SE Version 15.

## Results

Box 2 shows that 3.2% (n=5,646) of all adolescents aged 12-17 had a mental health-related hospitalisation, and 15.5% (n=27,203/175,115) of all adolescents had a child protection history. The proportion of adolescents with a mental health-related hospitalisation increased as the level of child protection contact increased, from 2.3% of those with no contact, to 6.0% of those with a notification, and 17.5% of individuals who had experienced out-of-home care. This pattern was consistent across all categories of mental health diagnoses. For example, for mental and behavioural disorders related to substance use, 0.8% of adolescents with no child protection contact had a hospitalisation compared to 1.9% of those with a notification and 9.6% of those in out-of-home care. Similarly, for self-harm, 0.7% of adolescents with no child protection contact had a hospitalisation compared to 2.1% of those with a notification, and 5.8% of those in out-of-home care. Supplementary Table 2 shows 32.4% of adolescents hospitalised for mental health had recurrent mental health-related hospitalisations. The proportion of adolescents with a recurrent hospitalisation increased as the level of child protection contact increased, from 28.6% of those with no contact, to 34.8% of those with a notification and 49.3% of those who had experienced out-of-home care. Overall, 74.9% of mental health-related hospitalisations were transferred from casualty/emergency departments (Supplementary Table 3).

#### Box 2: Proportion of individuals born in South Australia 1991-1999 (n=175,115) with at least one mental health-related hospitalisation at age 12-17 years, by highest level of contact with Child Protection (ages 0-11)

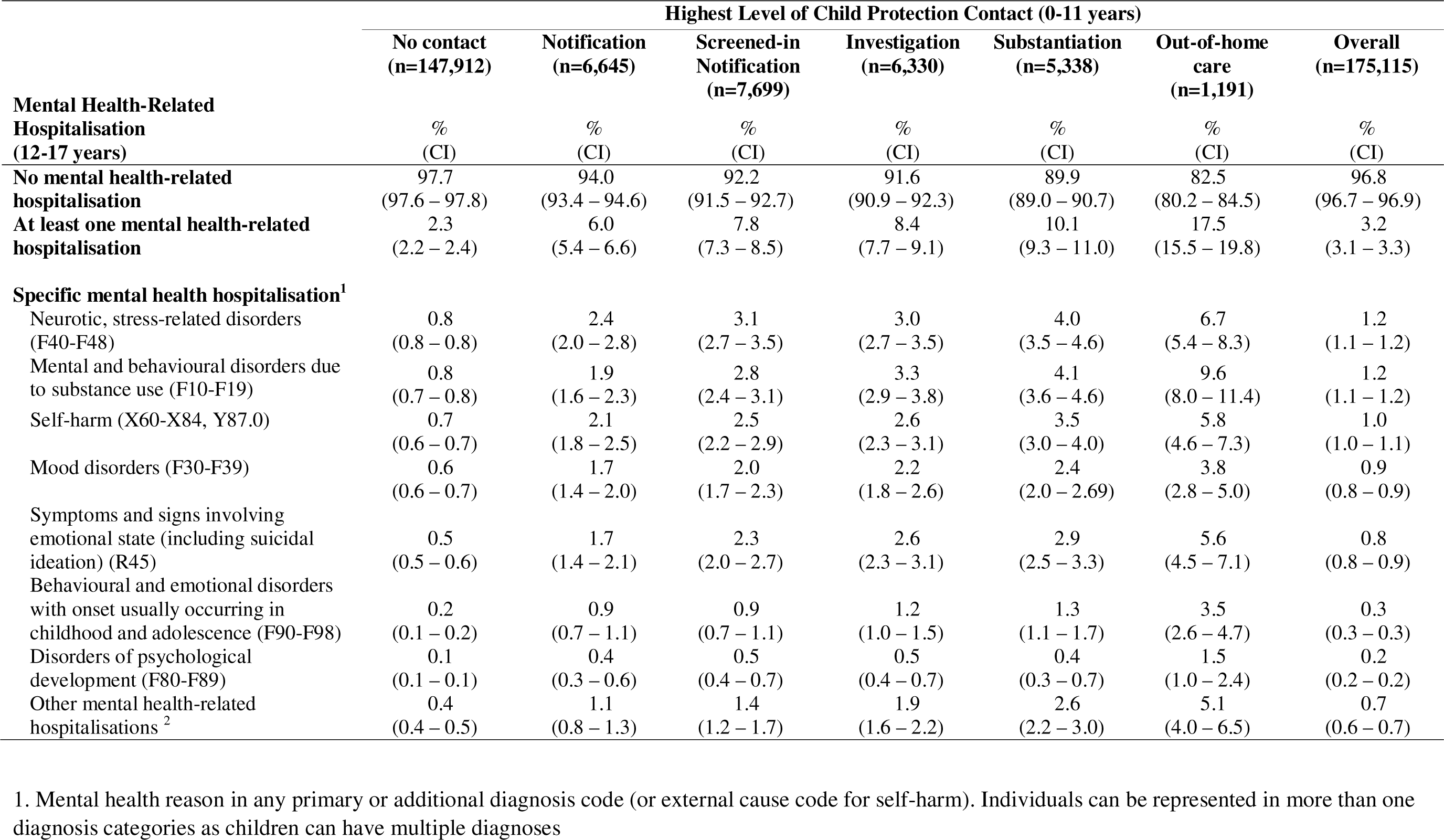
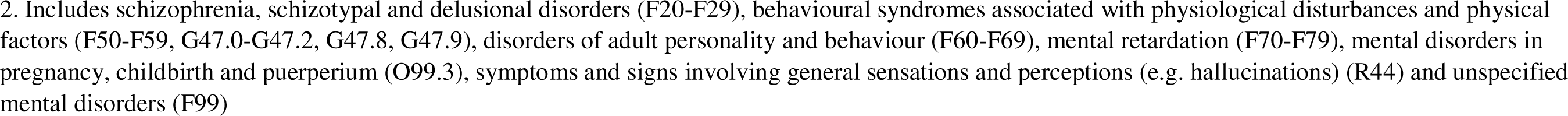

|  | Highest Level of Child Protection Contact (0-11 years) |  |  |  |  |  | Overall<br>(n=175,115) |
| --- | --- | --- | --- | --- | --- | --- | --- |
|  | No contact<br>(n=147,912) | Notification<br>(n=6,645) | Screened-in<br>Notification<br>(n=7,699) | Investigation<br>(n=6,330) | Substantiation<br>(n=5,338) | Out-of-home<br>care<br>(n=1,191) |  |
| **Mental Health-Related**<br>**Hospitalisation**<br>**(12-17 years)** | %<br>(CI) | %<br>(CI) | %<br>(CI) | %<br>(CI) | %<br>(CI) | %<br>(CI) | %<br>(CI) |
| **No mental health-related**<br>**hospitalisation** | 97.7<br>(97.6 – 97.8) | 94.0<br>(93.4 – 94.6) | 92.2<br>(91.5 – 92.7) | 91.6<br>(90.9 – 92.3) | 89.9<br>(89.0 – 90.7) | 82.5<br>(80.2 – 84.5) | 96.8<br>(96.7 – 96.9) |
| **At least one mental health-related**<br>**hospitalisation** | 2.3<br>(2.2 – 2.4) | 6.0<br>(5.4 – 6.6) | 7.8<br>(7.3 – 8.5) | 8.4<br>(7.7 – 9.1) | 10.1<br>(9.3 – 11.0) | 17.5<br>(15.5 – 19.8) | 3.2<br>(3.1 – 3.3) |
| **Specific mental health hospitalisation<sup>1</sup>** |  |  |  |  |  |  |  |
| Neurotic, stress-related disorders<br>(F40-F48) | 0.8<br>(0.8 – 0.8) | 2.4<br>(2.0 – 2.8) | 3.1<br>(2.7 – 3.5) | 3.0<br>(2.7 – 3.5) | 4.0<br>(3.5 – 4.6) | 6.7<br>(5.4 – 8.3) | 1.2<br>(1.1 – 1.2) |
| Mental and behavioural disorders due<br>to substance use (F10-F19) | 0.8<br>(0.7 – 0.8) | 1.9<br>(1.6 – 2.3) | 2.8<br>(2.4 – 3.1) | 3.3<br>(2.9 – 3.8) | 4.1<br>(3.6 – 4.6) | 9.6<br>(8.0 – 11.4) | 1.2<br>(1.1 – 1.2) |
| Self-harm (X60-X84, Y87.0) | 0.7<br>(0.6 – 0.7) | 2.1<br>(1.8 – 2.5) | 2.5<br>(2.2 – 2.9) | 2.6<br>(2.3 – 3.1) | 3.5<br>(3.0 – 4.0) | 5.8<br>(4.6 – 7.3) | 1.0<br>(1.0 – 1.1) |
| Mood disorders (F30-F39) | 0.6<br>(0.6 – 0.7) | 1.7<br>(1.4 – 2.0) | 2.0<br>(1.7 – 2.3) | 2.2<br>(1.8 – 2.6) | 2.4<br>(2.0 – 2.69) | 3.8<br>(2.8 – 5.0) | 0.9<br>(0.8 – 0.9) |
| Symptoms and signs involving<br>emotional state (including suicidal<br>ideation) (R45) | 0.5<br>(0.5 – 0.6) | 1.7<br>(1.4 – 2.1) | 2.3<br>(2.0 – 2.7) | 2.6<br>(2.3 – 3.1) | 2.9<br>(2.5 – 3.3) | 5.6<br>(4.5 – 7.1) | 0.8<br>(0.8 – 0.9) |
| Behavioural and emotional disorders<br>with onset usually occurring in<br>childhood and adolescence (F90-F98) | 0.2<br>(0.1 – 0.2) | 0.9<br>(0.7 – 1.1) | 0.9<br>(0.7 – 1.1) | 1.2<br>(1.0 – 1.5) | 1.3<br>(1.1 – 1.7) | 3.5<br>(2.6 – 4.7) | 0.3<br>(0.3 – 0.3) |
| Disorders of psychological<br>development (F80-F89) | 0.1<br>(0.1 – 0.1) | 0.4<br>(0.3 – 0.6) | 0.5<br>(0.4 – 0.7) | 0.5<br>(0.4 – 0.7) | 0.4<br>(0.3 – 0.7) | 1.5<br>(1.0 – 2.4) | 0.2<br>(0.2 – 0.2) |
| Other mental health-related<br>hospitalisations <sup>2</sup> | 0.4<br>(0.4 – 0.5) | 1.1<br>(0.8 – 1.3) | 1.4<br>(1.2 – 1.7) | 1.9<br>(1.6 – 2.2) | 2.6<br>(2.2 – 3.0) | 5.1<br>(4.0 – 6.5) | 0.7<br>(0.6 – 0.7) |

1. Mental health reason in any primary or additional diagnosis code (or external cause code for self-harm). Individuals can be represented in more than one diagnosis categories as children can have multiple diagnoses
2. Includes schizophrenia, schizotypal and delusional disorders (F20-F29), behavioural syndromes associated with physiological disturbances and physical factors (F50-F59, G47.0-G47.2, G47.8, G47.9), disorders of adult personality and behaviour (F60-F69), mental retardation (F70-F79), mental disorders in pregnancy, childbirth and puerperium (O99.3), symptoms and signs involving general sensations and perceptions (e.g. hallucinations) (R44) and unspecified mental disorders (F99)

We examined mental health hospitalisations in terms of the number of adolescents with at least one hospitalisation (person view), and the number of hospitalisations (hospital view). Box 3 shows there were 5,646 adolescents who experienced a total of 10,633 mental health-related hospitalisations from ages 12-17. Taking a person view, we see that although adolescents with a child protection history comprised only 15.5% of the population of 12-17-year-olds (Box 2), they made up 40.4% of the 12-17-year-olds with at least one mental health hospitalisation (Box 3). Taking a hospitalisations view: 44.9% of all mental health-related hospitalisations for 12-17-year-olds were for adolescents with a history of child protection contact (Box 3); adolescents who had been substantiated for child maltreatment by age 11 but were not in out-of-home care, accounted for 12.1% of all adolescent mental health-related hospitalisations, while those who had experienced out-of-home care from ages 0-11 accounted for 5.3% of all adolescent mental health-related hospitalisations.

#### Box 3: Distribution of individuals with at least one mental health-related hospitalisation, and mental health-related hospitalisations, by highest level of child protection system contact (0-11 years)

| **Highest Level of Child Protection System Contact** | **Adolescents** |  | **Hospitalisations** |  |
| --- | --- | --- | --- | --- |
|  | **Adolescents with at least one mental health-related hospitalisation** |  | **Hospitalisations that were mental health-related** |  |
|  | n children | % of all children | n MH hospitalisations | % of all MH hospitalisations |
| No contact | 3366 | 59.6 | 5856 | 55.1 |
| Any child protection contact | 2280 | 40.4 | 4777 | 44.9 |
| *Notification* | 398 | 7.0 | 776 | 7.3 |
| *Screened-in notification* | 603 | 10.7 | 1132 | 10.6 |
| *Investigation* | 530 | 9.4 | 1016 | 9.6 |
| *Substantiation* | 540 | 9.6 | 1285 | 12.1 |
| *Out-of-home care* | 209 | 3.7 | 568 | 5.3 |
| Total | 5646 | 100.0 | 10633 | 100.0 |

Box 4 takes a person view and shows that for those who had been hospitalised for any cause from ages 12-17, 14.9% had at least one mental health hospitalisation. The proportion of adolescents hospitalised for mental health, out of all adolescents hospitalised for any reason, increased with each level of child protection contact and was 3.3 times higher for adolescents who had experienced out-of-home care (39.1%) than adolescents with no contact (11.8%, Supplementary Table 4). This trend is consistent for each age, but is starkest at younger ages. For instance, for 17-year-olds the proportion with a mental health hospitalisation out of those hospitalised for any reason is 2.3 times higher for those who experienced out-of-home care compared to those with no child protection contact. However, the proportion with a mental health hospitalisation out of those hospitalised for any reason was 9.3 times higher for 12-year-olds with experience in out-of-home care than for 12-year-olds with no child protection system contact (Supplementary Table 4). This indicates that mental health is the reason for hospitalisation for a higher proportion of younger adolescents with out-of-home care experience than older adolescents with out-of-home care experience. Box 5 takes a hospitalisation view and shows the same pattern, with mental health diagnoses responsible for an increasing number of hospitalisations among younger adolescents and those with child protection contact.

#### Box 4: Person View: Proportion of children with at least one hospitalisation who had been hospitalised for a mental health (MH) condition by age (12-17 years) and highest level of child protection system contact^1^ (0-11 years)

| Highest Level of Child Protection System Contact |  |  |  |  |  |  |  |  |  |  |  |  |  |  |
| --- | --- | --- | --- | --- | --- | --- | --- | --- | --- | --- | --- | --- | --- | --- |
|  | No contact |  | Notification |  | Screened-in notification |  | Investigation |  | Substantiation |  | Out-of-home care |  | Overall |  |
| Age | n with MH hospitalisation / n with all cause hospitalisation | % MH of all hospitalised | n with MH hospitalisation / n with all cause hospitalisation | % MH of all hospitalised | n with MH hospitalisation / n with all cause hospitalisation | % MH of all hospitalised | n with MH hospitalisation / n with all cause hospitalisation | % MH of all hospitalised | n with MH hospitalisation / n with all cause hospitalisation | % MH of all hospitalised | n with MH hospitalisation / n with all cause hospitalisation | % MH of all hospitalised | n with MH hospitalisation / n with all cause hospitalisation | % MH of all hospitalised |
| 12 | 104/4857 | 2.1 | 12/317 | 3.8 | 29/437 | 6.6 | 29/398 | 7.3 | 33/342 | 9.6 | 19/97 | 19.6 | 226/6448 | 3.5 |
| 13 | 210/5064 | 4.1 | 32/390 | 8.2 | 49/468 | 10.5 | 63/392 | 16.1 | 67/411 | 16.3 | 23/112 | 20.5 | 444/6837 | 6.5 |
| 14 | 515/5900 | 8.7 | 57/411 | 13.9 | 110/559 | 19.7 | 98/467 | 21.0 | 104/477 | 21.8 | 47/138 | 34.1 | 931/7952 | 11.7 |
| 15 | 845/6747 | 12.5 | 108/530 | 20.4 | 165/665 | 24.8 | 151/574 | 26.3 | 146/517 | 28.2 | 58/143 | 40.6 | 1473/9176 | 16.1 |
| 16 | 1052/7442 | 14.1 | 145/608 | 23.8 | 200/782 | 25.6 | 164/678 | 24.2 | 189/649 | 29.1 | 92/184 | 50.0 | 1842/10343 | 17.8 |
| 17 | 1303/8309 | 15.7 | 144/662 | 21.8 | 222/891 | 24.9 | 192/761 | 25.2 | 201/720 | 27.9 | 68/186 | 36.6 | 2130/11529 | 18.5 |
| 12-17 | 3366/28629 | 11.8 | 398/2044 | 19.5 | 603/2522 | 23.9 | 530/2130 | 24.9 | 540/1966 | 27.5 | 209/534 | 39.1 | 5646/37825 | 14.9 |

1. Individuals in this table can have multiple hospitalisations at multiple ages and therefore may appear more than once

#### Box 5: Hospital View: Proportion of all hospitalisations due to mental health (MH) by highest level of child protection system contact and age

| Highest Level of Child Protection System Contact |  |  |  |  |  |  |  |  |  |  |  |  |  |  |
| --- | --- | --- | --- | --- | --- | --- | --- | --- | --- | --- | --- | --- | --- | --- |
|  | No contact |  | Notification |  | Screened-in notification |  | Investigation |  | Substantiation |  | Out-of-home care |  | Overall |  |
| Age | n MH hospitalisations / n all cause hospitalisations | % MH of all cause | n MH hospitalisations / n all cause hospitalisations | % MH of all cause | n MH hospitalisations / n all cause hospitalisations | % MH of all cause | n MH hospitalisations / n all cause hospitalisations | % MH of all cause | n MH hospitalisations / n all cause hospitalisations | % MH of all cause | n MH hospitalisations / n all cause hospitalisations | % MH of all cause | n MH hospitalisations / n all cause hospitalisations | % MH of all cause |
| 12 | 128/6607 | 1.9 | 15/418 | 3.6 | 33/638 | 5.2 | 42/600 | 7.0 | 47/488 | 9.6 | 23/131 | 17.6 | 288/8882 | 3.2 |
| 13 | 301/6943 | 4.3 | 39/527 | 7.4 | 74/690 | 10.7 | 87/583 | 14.9 | 93/564 | 16.5 | 34/170 | 20.0 | 628/9477 | 6.6 |
| 14 | 721/8310 | 8.7 | 78/585 | 13.3 | 173/826 | 20.9 | 132/765 | 17.3 | 193/710 | 27.2 | 90/221 | 40.7 | 1387/11417 | 12.1 |
| 15 | 1235/9490 | 13.0 | 156/748 | 20.9 | 223/958 | 23.3 | 204/921 | 22.2 | 265/788 | 33.6 | 137/286 | 47.9 | 2220/13191 | 16.8 |
| 16 | 1565/11045 | 14.2 | 219/926 | 23.7 | 276/1146 | 24.1 | 237/1046 | 22.7 | 327/1053 | 31.1 | 156/317 | 49.2 | 2780/15533 | 17.9 |
| 17 | 1906/12346 | 15.4 | 269/1063 | 25.3 | 353/1424 | 24.8 | 314/1205 | 26.1 | 360/1195 | 30.1 | 128/312 | 41.0 | 3330/17545 | 19.0 |
| 12-17 | 5856/54741 | 10.7 | 776/4267 | 18.2 | 1132/5682 | 19.9 | 1016/5120 | 19.8 | 1285/4798 | 26.8 | 568/1437 | 39.5 | 10633/76045 | 14.0 |

## Discussion

Children with child protection system contact from ages 0-11 accounted for 44.9% of all mental health-related hospitalisations among adolescents aged 12-17. Children with confirmed maltreatment (substantiated or out-of-home care) accounted for 17.4% of all adolescent mental health-related hospitalisations even though they comprised only 3.7% of the population aged 12-17. Among adolescents aged 12-17 who had at least one mental health-related hospitalisation, two in every five (40.4%) had previous child protection contact.

Our estimate that 3.2% of all adolescents had experienced a mental health-related hospitalisation was consistent with the Second Australian Child and Adolescent Survey of Mental Health and Wellbeing that found 3.3% of 12-17-year-olds had a mental disorder that severely impacted social and educational functioning in 2013-2014.^2^ Our study demonstrated that the out-of-home care population had the highest risk of mental health-related hospitalisations, with 17.5% of children who had experienced out-of-home care having at least one mental health-related hospitalisation in adolescence, which was consistent with previous Australian studies.^14, 15^ From the perspective of the hospital system, the proportion of all hospitalisations for adolescents aged 12-17 years that were mental health-related was higher for children who had experienced out-of-home care (39.5%) compared to adolescents with no child protection contact (10.7%). This comparison was more pronounced at younger ages, with the proportion of hospitalisations due to mental health being 9.3 times higher for 12-year-olds who experienced out-of-home care than for 12-year-olds with no child protection contact.

In 2020-21, SA Health and the Department for Child Protection signed an agreement reaffirming the prioritisation of health services for children in out-of-home care, including mental health services.^20^ However, we know that children who most need mental health support are more likely to be turned away from services due to their complex needs that services cannot meet.^21^ The Royal Australasian College of Physicians position statement on the Health Care of Children in Care and Protection Services highlighted the need for specialist multidisciplinary services for “vulnerable” children and adolescents to deliver integrated mental health, primary and other specialised care, as well as the need for trauma-informed health care for children and adolescents in out-of-home care.^22^

While this study indicates that priority access to mental health services is needed for children in out-of-home care, 39.6% of mental health-related hospitalisations are for adolescents who have had child protection system contact, but have not experienced out-of-home care. Even children who had only ever been notified to child protection were more than twice as likely to have a mental health hospitalisation. They also had a greater number of hospitalisations compared to those with no child protection contact. If mental health-related hospitalisations are to be reduced, mental health support services need to consider the heightened mental health needs of all children with child protection system contact, not just those in out-of-home care.^21^ As a broader system, we must also consider capacity to holistically address broader determinants of mental health such as poverty.^23, 24^ From a public health perspective, this means considering how we can provide early support before circumstances result in a mental health hospitalisation. Given the results of this study, child protection concerns may themselves be an indicator of need for early support.

This study excluded children not born in South Australia, although we would not expect the observed patterns identified to differ based on this. Additionally, private hospital data were not available. However, children in out-of-home care are unlikely to have private health cover unless it is funded by their carers.^25^ This means that most children in out-of-home care rely on public health services and most mental health-related hospitalisations among this group would have been captured in this analysis.^25^ This study also underestimated the total population burden of mental health challenges as we only use mental health-related hospitalisations as our measure of mental health challenges experienced by adolescents. However, given the similar magnitude of child protection contact across the life course in jurisdictions with different legislation and practice^12^ and the similar burden of mental health hospitalisations,^8^ we expect these findings to be applicable across Australia.

This paper provides evidence of the cross-over between child protection and subsequent mental health-related hospitalisations that reflects what clinicians are experiencing in hospitals across Australia. Almost 45% of mental health-related hospitalisations are experienced by adolescents who had prior child protection system contact. Over 40% of adolescents hospitalised for mental health had previous child protection system contact. As this child protection contact is prior to the subsequent mental health-related hospitalisations, this presents an opportunity for prevention of mental health challenges and hospitalisations among adolescents.

## Data Availability

All data analysed in the present study are not available.

