## Supplementary Tables for "Mental health-related hospitalisations among adolescents with previous child protection contact from birth to age 11"

**Supplementary Table 1:** ICD-10-AM codes used for mental health-related hospitalisations

| **Condition/Category of Conditions** | **ICD-10-AM Code/s** |
| --- | --- |
| Mental and behavioural disorders due to psychoactive substance use | F10-F19 |
| Mood disorders (e.g. depression) | F30-F39 |
| Neurotic, stress-related and somatoform disorders | F40-F48 |
| Disorders of psychological development (e.g. Autism) | F80-F89 |
| Behavioural and emotional disorders with onset usually occurring in childhood and adolescence | F90-98 |
| Signs and symptoms involving emotional state (e.g. nervousness, demoralisation and apathy) | R45 |
| Intentional self-harm^1^ | X60-X84, Y87.0 |
| Other^2^ | F20-F29, F50-59, G47.0-G47.2, G47.8, G47.9, F60-F69, F70-F79, O99.3, R44, F99 |

1. These are external cause codes.

2. Includes schizophrenia, schizotypal and delusional disorders, behavioural syndromes associated with physiological disturbances and physical factors (e.g. sleep disorders and eating disorders), disorders of adult personality and behavior, mental retardation, mental disorders in pregnancy, childbirth and puerperium, symptoms and signs involving general sensations and perceptions (e.g. auditory and visual hallucinations) and unspecified mental disorders.

**Supplementary Table 2:** Number of recurrent mental health-related hospitalisations for adolescents with at least one mental health-related hospitalisation (12-17) by highest level of child protection contact (0-11)

|  | **Highest Level of Child Protection System Contact** | | | | | | | | | | | | | |
| --- | --- | --- | --- | --- | --- | --- | --- | --- | --- | --- | --- | --- | --- | --- |
| **Number of recurrent mental health-related hospitalisations** | **No contact** | | **Notification** | | **Screened-in notification** | | **Investigation** | | **Substantiation** | | **Out-of-home care** | | **Overall** | |
|  | n | % | n | % | n | % | n | % | n | % | n | % | n | % |
| No recurrent hospitalisations | 2406 | 71.5 | 259 | 65.1 | 389 | 64.5 | 340 | 64.2 | 319 | 59.1 | 106 | 50.7 | 3819 | 67.6 |
| 1 recurrent hospitalisation | 531 | 15.8 | 71 | 17.8 | 124 | 20.6 | 98 | 18.5 | 114 | 21.1 | 46 | 22.0 | 984 | 17.4 |
| 2 recurrent hospitalisations | 177 | 5.3 | 28 | 7.0 | 36 | 6.0 | 44 | 8.3 | 42 | 7.8 | 22 | 10.5 | 349 | 6.2 |
| 3 recurrent hospitalisations | 91 | 2.7 | 10 | 2.5 | 18 | 3.0 | 17 | 3.2 | 27 | 5.0 | 11 | 5.3 | 174 | 3.1 |
| 4 or more recurrent hospitalisations | 161 | 4.8 | 30 | 7.5 | 36 | 6.0 | 31 | 5.8 | 38 | 7.0 | 24 | 11.5 | 320 | 5.7 |
| **Total** | 3366 | 100.0 | 398 | 100.0 | 603 | 100.0 | 530 | 100.0 | 540 | 100.0 | 209 | 100.0 | 5646 | 100.0 |

**Supplementary Table 3:** Where the patient was referred or transferred from for the mental health-related hospitalisation (12-17) by level of child protection contact (0-11)

|  | **Highest Level of Child Protection System Contact** | | | | | | | | | | | | | |
| --- | --- | --- | --- | --- | --- | --- | --- | --- | --- | --- | --- | --- | --- | --- |
| **Where the patient was referred/transferred from for admission to hospital** | **No contact** | | **Notification** | | **Screened-in notification** | | **Investigation** | | **Substantiation** | | **Out-of-home care** | | **Overall** | |
|  | n | % | n | % | n | % | n | % | n | % | n | % | n | % |
| Private medical practice | 314 | 5.4 | 36 | 4.6 | 64 | 5.7 | 65 | 6.4 | 56 | 4.4 | 13 | 2.3 | 548 | 5.2 |
| Inter Hospital Transfer | 145 | 2.5 | 30 | 3.9 | 38 | 3.4 | 30 | 3.0 | 50 | 3.9 | 14 | 2.5 | 307 | 2.9 |
| Outpatient Department | 923 | 15.8 | 76 | 9.8 | 125 | 11.0 | 77 | 7.6 | 141 | 11.0 | 47 | 8.3 | 1389 | 13.1 |
| Casualty/Emergency | 4256 | 72.7 | 608 | 78.4 | 832 | 73.5 | 798 | 78.5 | 998 | 77.7 | 473 | 83.3 | 7965 | 74.9 |
| Other^1^ | 218 | 3.7 | 26 | 3.4 | 73 | 6.4 | 46 | 4.5 | 40 | 3.1 | 21 | 3.7 | 424 | 4.0 |
| Total | 5856 | 100.0 | 776 | 100.0 | 1132 | 100.0 | 1016 | 100.0 | 1285 | 100.0 | 568 | 100.0 | 10633 | 100.0 |

1. Includes community health services, administrative admission, law enforcement agency, private psychiatric practice, retrieval and other

**Supplementary Table 4:** Person View: Proportion of children with at least one hospitalisation who had been hospitalised for a mental health (MH) condition by age (12-17 years) and highest level of child protection system contact with prevalence ratios^1^

| **Highest Level of Child Protection System Contact** | | | | | | | | | | | |
| --- | --- | --- | --- | --- | --- | --- | --- | --- | --- | --- | --- |
|  | **No contact** | **Notification** | | **Screened-in notification** | | **Investigation** | | **Substantiation** | | **Out-of-home care** | |
| Age | % MH of all hospitalised  (Ref) | % MH of all hospitalised | MH hospitalised ratio | % MH of all hospitalised | MH hospitalised ratio | % MH of all hospitalised | MH hospitalised ratio | % MH of all hospitalised | MH hospitalised ratio | % MH of all hospitalised | MH hospitalised ratio |
| 12 | 2.1 | 3.8 | 1.8 | 6.6 | 3.1 | 7.3 | 3.5 | 9.6 | 4.6 | 19.6 | 9.3 |
| 13 | 4.1 | 8.2 | 2.0 | 10.5 | 2.6 | 16.1 | 3.9 | 16.3 | 4.0 | 20.5 | 5.0 |
| 14 | 8.7 | 13.9 | 1.6 | 19.7 | 2.3 | 21.0 | 2.4 | 21.8 | 2.5 | 34.1 | 3.9 |
| 15 | 12.5 | 20.4 | 1.6 | 24.8 | 2.0 | 26.3 | 2.1 | 28.2 | 2.3 | 40.6 | 3.2 |
| 16 | 14.1 | 23.8 | 1.7 | 25.6 | 1.8 | 24.2 | 1.7 | 29.1 | 2.1 | 50.0 | 3.5 |
| 17 | 15.7 | 21.8 | 1.4 | 24.9 | 1.6 | 25.2 | 1.6 | 27.9 | 1.8 | 36.6 | 2.3 |
| Total | 11.8 | 19.5 | 1.7 | 23.9 | 2.0 | 24.9 | 2.1 | 27.5 | 2.3 | 39.1 | 3.3 |

1. Individuals in this table can have multiple hospitalisations at multiple ages and therefore may appear more than once

The proportion of 12-year-olds in the out-of-home care group with at least one hospitalisation who had been hospitalised due to mental health challenges was 9.3 times higher than for 12-year-olds with no contact. The proportion of 17-year-olds in the out-of-home care group with at least one hospitalisation who had been hospitalised due to mental health challenges was 2.3 times higher than for 17-year-olds with no contact.

**Supplementary Table 5:** Hospital View: Proportion of all hospitalisations due to mental health (MH) by highest level of child protection system contact and age with mental health hospitalisations ratios

| **Highest Level of Child Protection System Contact** | | | | | | | | | | | |
| --- | --- | --- | --- | --- | --- | --- | --- | --- | --- | --- | --- |
|  | **No contact** | **Notification** | | **Screened-in notification** | | **Investigation** | | **Substantiation** | | **Out-of-home care** | |
| Age | % MH of all cause  (Ref) | % MH of all cause | MH hospitalisations ratio | % MH of all cause | MH hospitalisations ratio | % MH of all cause | MH hospitalisations ratio | % MH of all cause | MH hospitalisations ratio | % MH of all cause | MH hospitalisations ratio |
| 12 | 1.9 | 3.6 | 1.9 | 5.2 | 2.7 | 7.0 | 3.7 | 9.6 | 5.1 | 17.6 | 9.3 |
| 13 | 4.3 | 7.4 | 1.7 | 10.7 | 2.5 | 14.9 | 3.5 | 16.5 | 3.8 | 20.0 | 4.7 |
| 14 | 8.7 | 13.3 | 1.5 | 20.9 | 2.4 | 17.3 | 2.0 | 27.2 | 3.1 | 40.7 | 4.7 |
| 15 | 13.0 | 20.9 | 1.6 | 23.3 | 1.8 | 22.2 | 1.7 | 33.6 | 2.6 | 47.9 | 3.7 |
| 16 | 14.2 | 23.7 | 1.7 | 24.1 | 1.7 | 22.7 | 1.6 | 31.1 | 2.2 | 49.2 | 3.5 |
| 17 | 15.4 | 25.3 | 1.6 | 24.8 | 1.6 | 26.1 | 1.7 | 30.1 | 2.0 | 41.0 | 2.7 |
| Total | 10.7 | 18.2 | 1.7 | 19.9 | 1.9 | 19.8 | 1.9 | 26.8 | 2.5 | 39.5 | 3.7 |

The proportion of hospitalisations due to mental health for 12-year-olds with out-of-home care experience was 9.3 times higher than for hospitalisations for 12-year-olds with no contact. The proportion of hospitalisations due to mental health for 17-year-olds with out-of-home care experience was 2.7 times higher than for hospitalisations for 17-year-olds with no contact.
